# COVID-19 pandemic: Impact of lockdown, contact and non-contact transmissions on infection dynamics

**DOI:** 10.1101/2020.04.04.20050328

**Authors:** Shovonlal Roy

**Affiliations:** Department of Geography and Environmental Science, University of Reading, Whiteknights, Reading RG6 6AB, U.K.

**Keywords:** COVID-19, SEIR models, Non-contact transmission, Contact transmission, Social distancing, Lockdown

## Abstract

COVID-19 coronavirus pandemic has virtually locked down the entire world of human population, and through its rapid and unstoppable spread COVID-19 has essentially compartmentalised the population merely into susceptible, exposed, infected and recovered classes. Adapting the classical epidemic modelling framework, two distinct routes of COVID-19 transmission are incorporated into a model: (a) direct person-to-person contact transmission, and (b) indirect airborne and fomites-driven transmission. The indirect non-contact transmission route needs to explored in models of COVID-19 spread, because evidences show that this route of transmission is entirely viable with hugely uncertain level of relative contribution. This theoretical study based on model simulations demonstrates the following: (1) Not incorporating indirect transmission route in the model leads to underestimation of the basic reproduction number, and hence will impact on the COVID-19 mitigation decisions; (2) Lockdown measures can suppress the primary infection peak, but will lead to a secondary peak whose relative strength and time of occurrence depend on the success and duration of the lockdown measures; (3) To make lockdown effective, a considerable level of reduction in both contact and non-contact transmission rates over a long period is required; (4) To bring down the infection cases below any hypothetical health-care capacity, reduction of non-contact transmission rate is key, and hence active measures should be taken to reduce non-contact transmission (e.g., extensive uses of areal and aerosol disinfectant in public spaces to improve contaminated surfaces and air); (5) Any premature withdrawal of lockdown following the sign of a brief retracement in the infection cases can backfire, and can lead to a quicker, sharper and higher secondary peak, due to reactivation of the two transmission routes. Based on these results, this study recommends that any exit policy from lockdown, should take into account the level of transmission reduction in both routes, the absolute scale of which will vary among countries depending on their health-service capacity, but should be computed using accurate time-series data on infection cases and transmission rates.

## Introduction

Over the last few months, COVID-19 has expanded across the world, and is now challenging the very existence of human population in many squares on the Earth. The spread of this infection was rapid, and no specific group of the population was essentially immune to this infection. Reported daily deaths suggest that the immune-response to this virus may vary among age groups and depend on underlying health conditions (CDC COVID-19 report, 2020). The victims range most widely ([WHO report, 2019](https://www.who.int/docs/default-source/coronaviruse/who-china-joint-mission-on-covid-19-final-report.pdfunderline)), from new borns (< 1 month) or even unborn to elderly (90 years+), and from chronic patients suffering from asthma, other lungs conditions, heart disease, to healthy athletes and healthy teenagers ([BBC report](http://helpx.adobe.com/acrobat/using/allow-or-block-links-internet.html?mv=product&mv2=acrobat), [MMWR, 2020](https://www.cdc.gov/mmwr/volumes/69/wr/mm6913e2.htm?s_cid=mm6913e2_w)).

Given these evidences, it is reasonable to assume that COVID-19 has essentially turned the entire human population into a few distinct compartments: the susceptible, the exposed, the infected and the recovered groups, which are continuously interacting with each other. Classical epidemiological modelling framework [1–3], therefore, should be capable of describing this pandemic dynamics, and predicting the impact of mitigation measures currently undertaken by Governments worldwide.

In this paper, a compartmental epidemiological model is simulated, taking into consideration the transmission routes for COVID-19 infection among the susceptible (S), exposed (E), infected (I) and recovered (R) classes of the population (see Appendix for model equations). The model builds on classical SIR models [1–3], but is extended to incorporate two transmission routes and mitigation measures such as lockdown and social distancing, which has been undertaken across countries. The epidemic model includes the following **two routes of transmission**:

1. Person-to-person transmission, which is widely used in epidemiological models [2], and has been recognised as the primary route for COVID-19 transmission e.g. [4]. Strength of the transmission is incorporated in the model through a person-to-person contact rate (henceforth referred to as **‘contact transmission’**).
2. Airborne transmission and transmission via fomites, which are indirect (not as a consequence of direct person-to-person contact) routes for COVID-19 spread, and which have been recognised as possible causes for infection spread [5, 6]. Strength of the indirect transmission from all external sources (henceforth referred to as **‘non-contact transmission’**) is incorporated in the model through a non-contact transmission rate that affects the exposed group of the population.

It is important to highlight that, compared to directcontact exposure, little is known about the impact and relative strength of the indirect route of transmission for COVID-19, due to insufficient data. This constraint imposes uncertainty in predicting and mitigating COVID-19 infection within the boundary of any country. Scientists have further warned that it could take long time to gather sufficient data on this route of infection, which may cost more lives ([Nature report](https://www.nature.com/articles/d41586-020-00974-w#ref-CR1)). The role of indirect transmission on the infection dynamics, therefore, has been less explored or unexplored in the context of COVID-19 modelling. In other epidemiological models, however, the non-contact route of infection has been explored previously e.g., [7, 8]. Here, non-contact transmission is included in an SEIR model, and its impact is explored in conjunction with mitigation measures. Further details on model equation, parameterisation and simulations methods are given in the appendix.

## Results and discussion

### Non-contact transmission and basic reproduction number

Basic reproduction number (*R*_0_), which is usually defined as the average number of new infections caused by a single infected individual in the susceptible population, depends on the transmission routes and rates. For the SEIR model under investigation, *R*_0_ was calculated from the next-generation matrix [9]. The magnitude of *R*_0_ obtained with person-to-person contact as the only route of transmission, appears to be an underestimation when non-contact transmission is active (Fig. 1).

**Figure 1:**
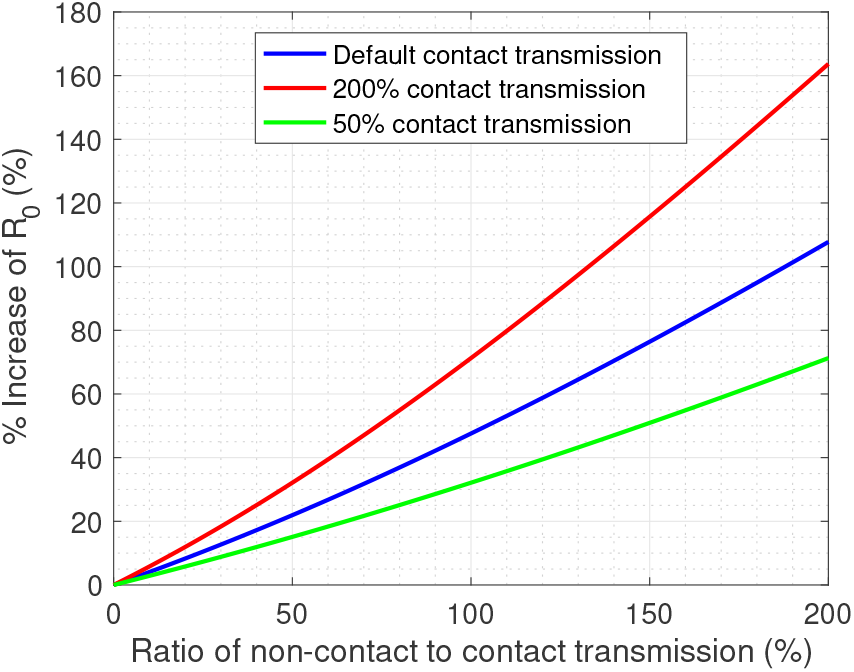
Change in *R*_0_ as a consequence of non-contact transmission. Shown are the changes in % *R*_0_ as a function of changes in non-contact to contact transmission rate. (*Disclaimer: In this figure relative (%) scales are used for the purpose of understanding the dynamics - time scale or absolute scales are not optimised to any Country’s data and so the scales shown may not be directly used in decision making.)*

If the relative contribution of non-contact transmission is very small, changes in *R*_0_ is generally negligible. But *R*_0_ increases sharply with the relative contribution of non-contact transmission (Fig. 1). For example, whilst a 50% relative contribution of non-contact transmission increases *R*_0_ by 15-35%, a 150% relative contribution can double it (depending on the nature of infection). Since in the case of COVID-19 the contribution of non-contact transmission is unknown, it is uncertain as to what extent *R*_0_ will spike if no action is taken to reduce this route of transmission.

### Non-contact transmission and impact of lockdown

An universal assumption behind lockdown measures is that the rate of infection will reduce, thereby suppressing the spread of infection. Unsurprisingly, the model simulations generally confirms this assumption (Fig. 2). Any level of reduction in transmission over a sufficiently-long lockdown period will lower the peak of infection from ‘no-action’ scenario (Fig. 2). The real success of lockdown measures, however, will be to keep the primary infection peak always below a Country’s health-services capacity (see, Fig. 2, the horizontal red lines representing some hypothetical health-service capacities). But this is possible only if the transmission rates reduce considerably - see Fig. 2 for various scenarios, and the decrease in peak as a consequence.

**Figure 2:**
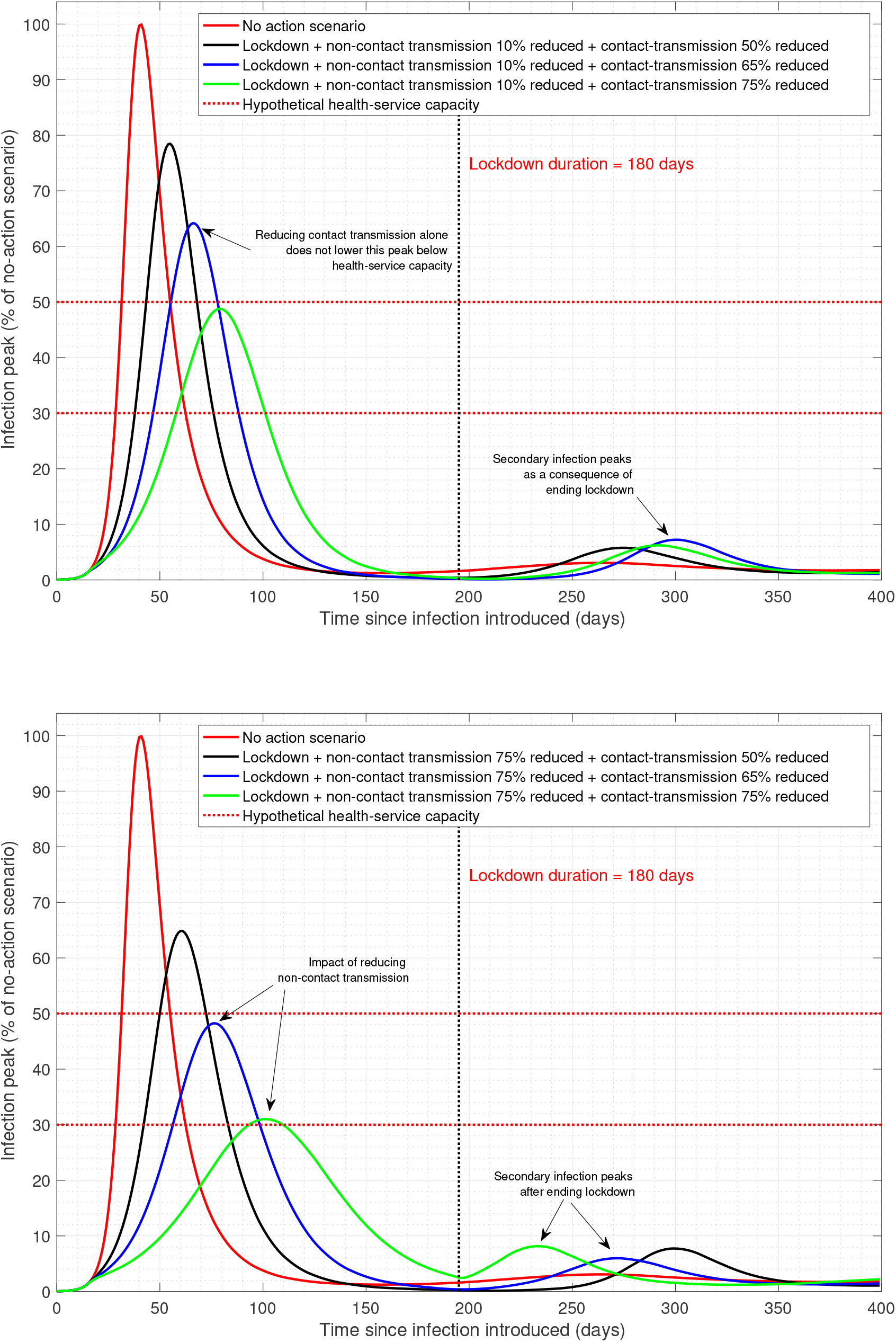
Impact of lockdown. Top panel - impact on infection dynamics when contact-transmission reduces greatly but non-contact transmission reduces slightly during lockdown. Bottom panel - impact on infection dynamics when both contact-transmission & non-contact transmission reduce significantly during lockdown. See supplementary for movie clips. (*Disclaimer: In this figure relative (%) scales are used for the purpose of understanding the dynamics - time scale or absolute scales are not optimised to any Country’s data and so the scales shown may not be directly used in decision making.)*

How far the transmission rates should reduce during lockdown will depend on the Country’s health service capacity - e.g., a 50% (on a relative scale) reduction of the infection peak obtained through 75% reduction in contact transmission during lockdown, may still be insufficient if the health-service capacity is only 30% (see, Fig. 2). Therefore, depending on the health-service capacity, stricter measures must be imposed during lock-down to ultimately bring down the infection peak to a level that can be handled at any given time. This result suggests that countries with poor health-service facility and capacity require stricter lockdown measures than those with higher capacity.

Simulations suggest that due consideration should be given to reduce not one, but all transmission rates, i.e both contact transmission and non-contact transmission. To give an example, let us suppose that the health-service capacity is only 30% of the no-action infection-peak. Under this constraint, if the contact transmission reduces by 75% during lockdown, but non-contact transmission does not reduce considerably (10%), the infection peak will stay well above the health-service capacity (top panel, green line in Fig. 2). But if the non-contact transmission further reduces, say, by 75%, the effect will be a considerably lower infection peak, within the reach of health-service capacity (bottom panel, green line in Fig. 2).

These results suggest that during lockdown, pro-active measures to reduce non-contact transmission, such as frequent and thorough sanitisation of public-access areas and wider uses of aerosol disinfectant, will be required to reduce the spread of infection and bring down the infection peak. Ignoring such measures may cause additional public distress, arising from far stricter lockdown measures, when mitigation strategy is solely based on contact route of transmission.

### Impact of ending lockdown prematurely

Whilst lockdown and associated measures guarantee initial reduction of the disease spread, any suppression of the infection peak results in a secondary infection peak after the lockdown has been lifted (Fig. 2). Theoretically, the secondary peak enforced by lockdown will be higher than that for the no-action scenario. It is, therefore, important to investigate the dynamics of the secondary peak for deciding the duration of lockdown (Fig. 3).

**Figure 3:**
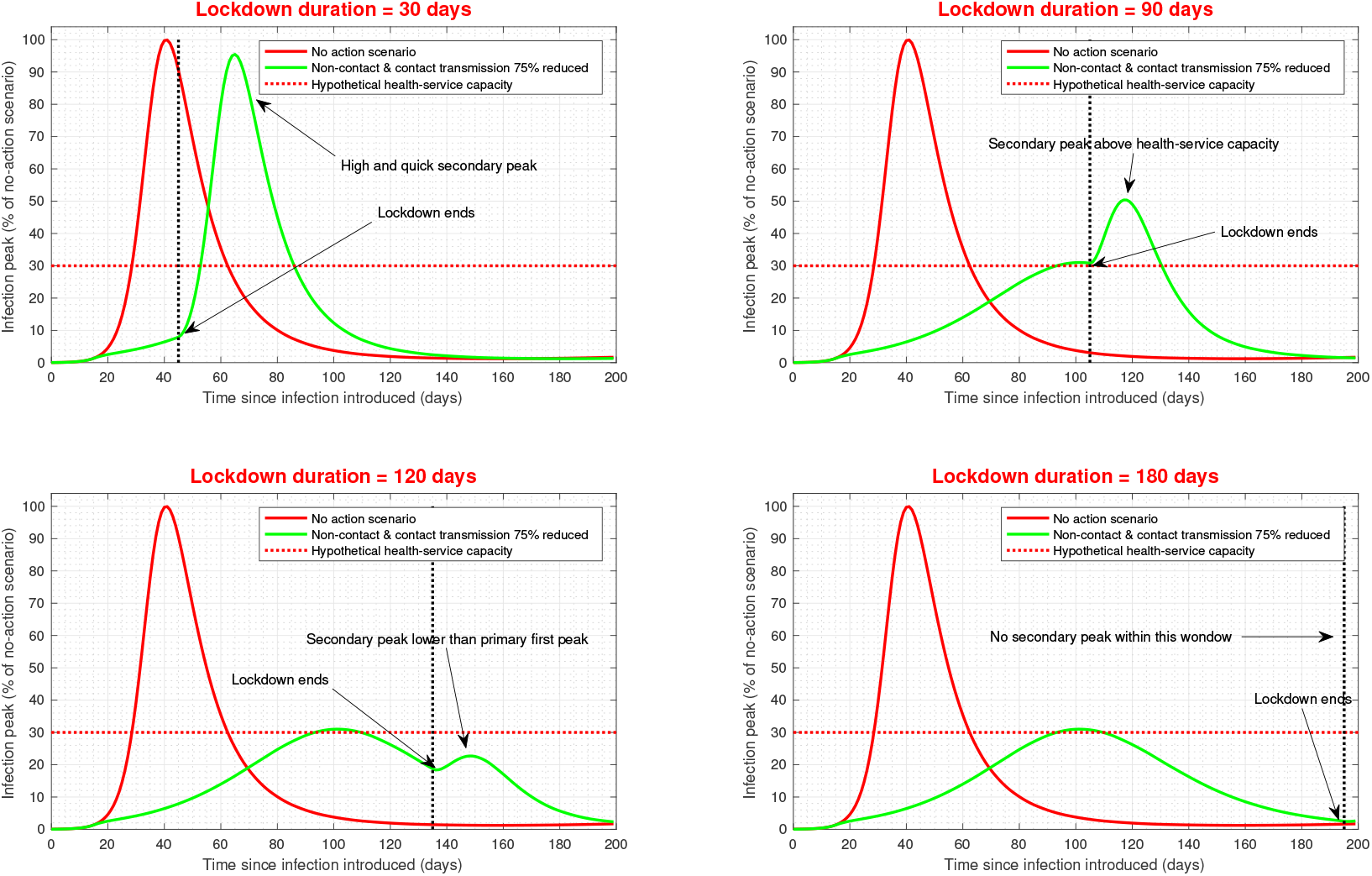
Impact of lockdown duration on primary and secondary peak dynamics. Evolution of primary and secondary peaks are shown under four hypothetical lockdown duration. See supplementary for a movie clip. (*Disclaimer: In this figure relative (%) scales are used for the purpose of understanding the dynamics - time scale or absolute scales are not optimised to any Country’s data and so the scales shown may not be directly used in decision making.)*

To demonstrate the secondary-peak dynamics, let us consider the best scenario out of those shown in (Fig. 2), i.e, both non-contact and contact transmission rates are reduced by 75% throughout the lockdown period. Under such an optimistic scenario, the evolution of secondary peak, its height, time of occurrence and duration solely depend on the duration of lockdown (see, green lines in Fig. 3). For example, with a hypothetical health-service capacity equal to 30% of the no-action peak, the first 30 days of lockdown show significant reduction in the infection cases (see, red and green lines in the first window, Fig. 3), giving a false impression that the subsequent infection cases will stay below the capacity. But once the lockdown is fully lifted, the infection cases spike rapidly over a short time, and the peak goes beyond the capacity, almost nearing the no-action peak. This scenario demonstrates the ultimate failure of the suppression measures as a consequence of premature end of lockdown.

When lockdown continues slightly longer, say, only until when the primary peak reduces down to the health-service capacity (e.g., 90 days in the second window, Fig. 3), the infection cases almost plateau, giving a false impression that a lower number of subsequent cases will follow beyond lockdown. But, after a minor retracement, the infection cases increase and the secondary peak goes beyond the health-service capacity, suggesting that this is also a premature end of lock-down.

When lockdown continues much longer, even after the infection cases plateau, and it continues until the cases are considerably below the health-service capacity (e.g., 120-180 days in window 3 and 4, Fig. 3), a characteristically different secondary peak evolves. The secondary peak is now lower than the primary peak, and hence much lower than the health-service capacity. Longer lockdown further pushes back secondary peak, giving adequate time for preparation for the second wave (Fig. 3).

### Non-contact and contact transmission and infection peaks after lockdown

With the infection suppression measures, it would be essential to check not only the primary peak, but also the secondary peak and the delay in peak occurrence under different transmission rates (Fig. 4). Although longer lockdown reduces the primary peak, the scale of the reduction does not change beyond a certain point of time (first and second window, Fig. 4). Non-contact transmission plays a key role here, and if this transmission rate is proactively reduced through disinfection measures, the impact of longer lockdown will be realised on the number of new cases. Under a hypothetical scenario, with a 50% reduction in contact transmission and 10% reduction in non-contact transmission, any impact on the primary peak and its occurrence may cease after 30 days of lockdown. However, under the same set up, if both transmissions reduce by 75%, the impact on primary peak and its occurrence can persist 90 days and beyond, which will bring down the primary infection cases (first and second window, Fig. 4).

**Figure 4:**
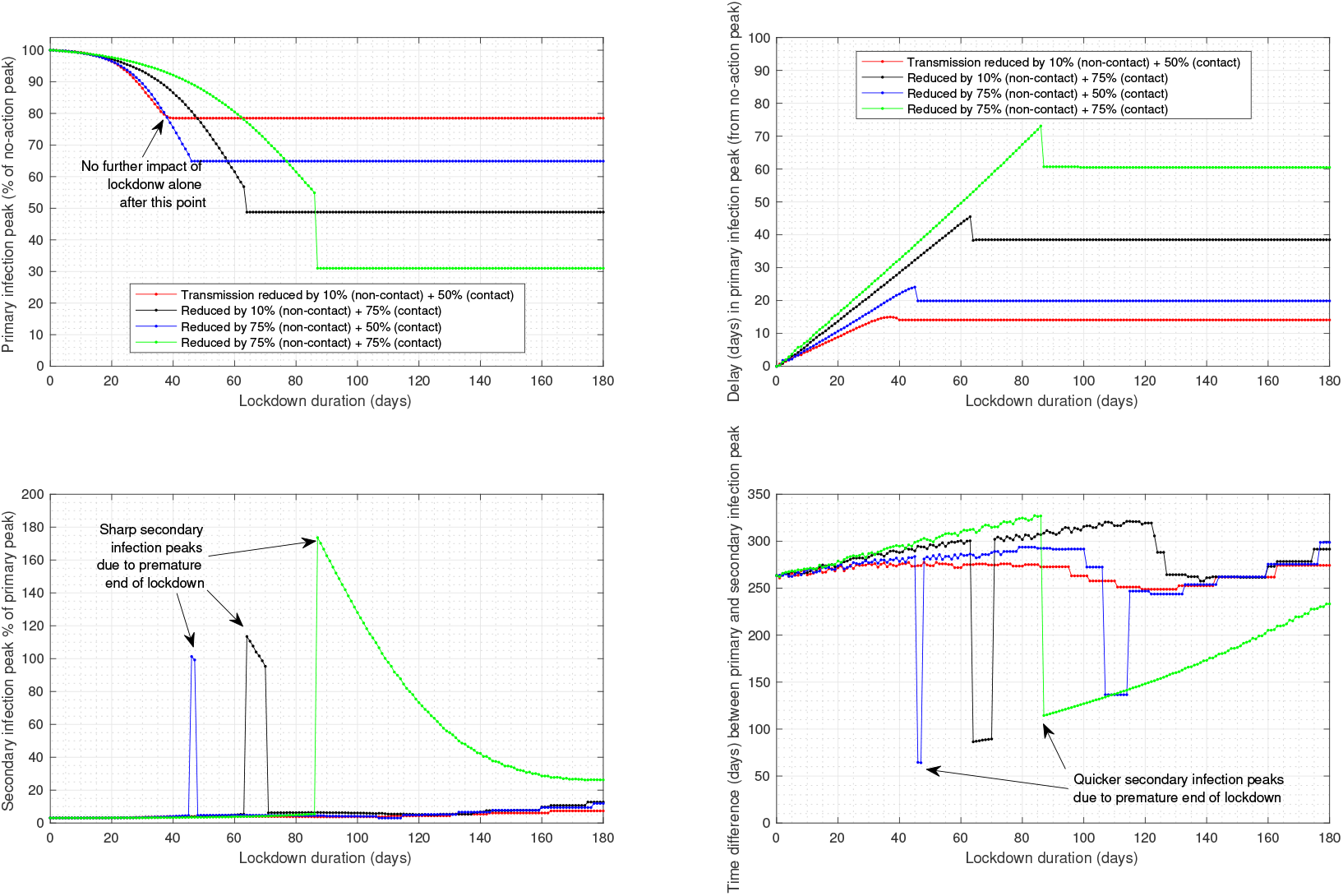
Infection peaks after lockdown: under different non-contact and contact transmission transmission and duration of lockdown. Top panels demonstrate the primary peak and delay in primary peak relative to the no-action scenario. Bottom panels demonstrate the primary peak and timing of secondary peak under various scenarios. (*Disclaimer: In this figure relative (%) scales are used for the purpose of understanding the dynamics - time scale or absolute scales are not optimised to any Country’s data and so the scales shown may not be directly used in decision making.)*

A reduced impact of lockdown on primary peak needs to be considered in conjunction with characteristics of the enforced secondary peak. For any level of reduction in contact and non-contact transmission, the secondary peak and its occurrence depend on the duration of lockdown measures (third and fourth window, Fig. 4). Under a hypothetical scenario, with a 75% reduction in contact transmission and 10% reduction in non-contact reduction, the secondary peak can be vigorous: higher than the primary peak, and occurring within 60 days of the first peak (black lines, in third and fourth window, Fig. 4).

## Concluding remarks

Based on a compartmental SEIR epidemic modelling framework the dynamics of an infected population is studied here. The model incorporates two different routes of COVID-19 transmission: the traditional person-to-person contact route, and the less explored fomites-mediated and airborne route. Whilst the preliminary evidence suggest that both transmissions contribute to COVID-19 spread [5, 6], the impact of non-contact transmission has been recognised as more uncertain due to difficulty in data collection, and therefore requires proper attention in modelling studies.

Indirect transmission via non-contact route can increase the basic reproduction number significantly. Therefore, not incorporating this route of transmission in the model will underestimate the strength of COVID-19 transmission and spread.

Simulation results suggest that lockdown will impact on infection spread, and initially suppress the number of infection cases to a great extent. But lockdown alone will not eradicate the infection from the system, particularly if the duration of lockdown is short, and if stricter measures are not taken to reduce all the routes of disease transmission.

Premature withdrawal of lockdown is likely to promote a rapid and sharper infection peak, which may result in more new infection cases than the capacity of national health-care services. Premature withdrawal of lockdown may be influenced by the ‘false signal’ of an initial plateau or even a minor dip in the number of cases, which is a trivial consequence of initial decrease in contact transmission. If the lockdown ends prematurely, a brief retracement may follow, but after this, the infection cases can in no time jump to a new high due to reactivation of the transmission routes.

During the lockdown period, strong measures will be required to significantly reduce the routes of transmission. Contact transmission should be reduced as much as possible. There is, however, the possibility that the indirect or non-contact transmission route is ignored, which will be less helpful in reducing the infection cases in situation where the relative strength of this transmission rate is high. Therefore, pro-active measures should be taken to reduce non-contact transmission. Some of those measures may involve frequent and wider uses of areal and aerosol disinfectant in public spaces to improve contaminated surfaces and air, as has been done by certain countries (e.g., China, [World Economic Forum report](https://www.weforum.org/agenda/2020/03/three-ways-china-is-using-drones-to-fight-coronavirus/)). Ignoring non-contact transmission should not be compensated by far stricter lockdown or unachievable lockdown measures, as the mitigation strategy should not be solely based on contact route of transmission.

Any exit policy from lockdown should carefully consider the level of success in reducing both the routes of transmission over a sufficiently long time frame. The absolute scale of the required measures such as, % reduction in both contact and non-contact transmission, will vary for countries depending on their health-service capacity, and could be computed from the model used in this paper using accurate time-series data on infection cases and transmission rates (which is beyond the scope of this paper). An optimal lockdown duration or a ‘sweet-spot’ for lockdown withdrawal, therefore, can differ between countries.

## Data Availability

The data presented are based on a model simulation; the model code is available upon request.

## Appendix

### Model, parametrisation and simulation

A compartmental susceptible(S)-exposed(E)-infected(I)-recovered(R) model [1–3] is adapted with two routes of COVID-19 transmission. Total population is *N* (*t*) = *S*(*t*) + *E*(*t*) + *I*(*t*) + *R*(*t*), and no further demographic parameters are included. Susceptible population is governed by the equation: 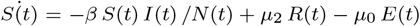 where*β* is the average person-to-person contact transmission rate day−1, *µ*_0_ is the average non-contact transmission day−1, and *µ*_2_ is the rate of recovered people converting into susceptibleday−1. Exposed population is governed by the equation: 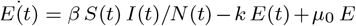; where *k* is the rate of converting from exposed to infected day−1. Infected population is governed by the equation: 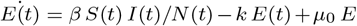; where *γ* is the recovery rate for the infected day−1, and *µ*_1_ is the death rate of the infected day−1. Finally, the recovered population is governed by the equation: 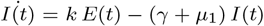.

A range of parameter values are included in the simulations, and the parameter ranged are taken from various COVID-19 studies published online. The parameter ranges are as follows: *β* = [0.12, 0.5], *k* = [0.2, 0.3], *γ* = [0.25, 0.35], *µ*_1_ = [0.025, 0.05], *µ*2 = [0, 0.005], *µ*_0_ = [0, 0.1]. All simulations are done using MATLAB R2016b package. The code will be available upon request.

